# Predictors of acute ischemic cerebral lesions in immune-mediated thrombotic thrombocytopenic purpura and hemolytic uremic syndrome

**DOI:** 10.1101/2023.04.05.23288216

**Authors:** Lisa Neuman, Adrien Joseph, Raïda Bouzid, Mickael Lescroart, Eric Mariotte, Stéphane Ederhy, Sophie Tuffet, Jean-Luc Baudel, Ygal Benhamou, Lionel Galicier, Steven Grangé, François Provôt, Antoine Neel, Frédéric Pène, Yahsou Delmas, Claire Presne, Pascale Poullin, Alain Wynckel, Pierre Perez, Christelle Barbet, Jean-Michel Halimi, Valérie Chatelet, Jean-Michel Rebibou, Mario Ojeda-Uribe, Cécile Vigneau, Laurent Mesnard, Agnès Veyradier, Elie Azoulay, Paul Coppo, Hugues Chabriat, the participants to the Prospective Multicenter MATRISK study

## Abstract

**Background:** The immune form of thrombotic thrombocytopenic purpura (iTTP) and the hemolytic and uremic syndrome (HUS) are two major forms of thrombotic microangiopathy (TMA). Their treatment has been recently greatly improved. In this new era, both the prevalence and predictors of cerebral lesions occurring during the acute phase of these severe conditions remain poorly known.

**Aim:** The prevalence and predictors of cerebral lesions appearing during the acute phase of iTTP and shigatoxin-producing *Escherichia coli*-HUS or atypical HUS were evaluated in a prospective multicenter study.

**Methods:** Univariate analysis was performed to report the main differences between patients with iTTP and those with HUS or between patients with acute cerebral lesions and the others. Multivariable logistic regression analysis was used to identify the potential predictors of these lesions.

**Results:** Among 73 TMA cases (mean age 46.89 ± 15.99 years (range: 21-87 years) with iTTP (n = 57) or HUS (n= 16), one third presented with acute ischemic cerebral lesions on magnetic resonance imagery (MRI); two individuals also had hemorrhagic lesions. One in ten patients had acute ischemic lesions without any neurological symptom. The neurological manifestations did not differ between iTTP and HUS. In multivariable analysis, 3 factors predicted the occurrence of acute ischemic lesions on cerebral MRI: 1) the presence of old infarcts on cerebral MRI, 2) the level of blood pulse pressure, 3) the diagnosis of iTTP.

**Conclusion:** Cerebral MRI is crucial to detect both symptomatic and covert ischemic lesions at the acute phase of iTTP and HUS and helps identify patients with old infarcts, at the highest risk of neurological worsening. The diagnosis of iTTP further increases the risk of ischemic lesions but also an increased level of blood pressure that may represent a potential target to further improve the therapeutic management of these conditions.

**Key points:**

- One third of patients with immune-mediated thrombotic thrombocytopenic purpura (iTTP) or the hemolytic uremic syndrome (HUS) present with acute ischemic cerebral lesions on magnetic resonance imagery (MRI);
- The cerebral manifestations on MRI did not differ between iTTP and HUS;
- The presence of old infarcts on cerebral MRI, the level of blood pulse pressure and a diagnosis of iTTP predict the occurrence of acute ischemic lesions on cerebral MRI.

## Introduction

Thrombotic thrombocytopenic purpura (TTP) and the Hemolytic and Uremic Syndrome (HUS) are two forms of thrombotic microangiopathy (TMA). Their clinical and biological presentation includes mechanical hemolytic anemia and consumptive thrombocytopenia with microthrombi that can lead to various ischemic end-organ damage^1^. A severe antibody-mediated deficiency of ADAMTS13, the metalloproteinase cleaving Willebrand factor^2^, is responsible for the immune form of TTP (iTTP). In contrast, HUS can result from <u>S</u>higa<u>t</u>oxin produced during infections by *Escherichia Coli* (Shigatoxin-associated *Escherichia coli* [STEC]-HUS) or from a deregulation of the complement alternative pathway (atypical HUS).^1^ These last decades, the short-term mortality of iTTP has been dramatically reduced with therapeutic plasma exchanges, corticosteroids, rituximab targeting B-cells, and caplacizumab targeting the von Willebrand factor^3^. Also, the prognosis of HUS has been considerably improved with standardized supportive care^4^ as well as anti-C5 monoclonal antibody (eculizumab) in atypical HUS.^5^

In this new therapeutic era, the prevalence of neurological complications and pattern of brain damage observed during these rare conditions remain poorly described both at the acute phase and during long-term follow-up. Recent data suggest that a significant proportion of survivors could present with symptomatic but also silent cerebral injuries^6^. In the present study, we aimed to report the brain lesions appearing in the acute phase of iTTP and HUS and whether predictors of these acute lesions could be identified. For this purpose, we determined the frequency and type of lesions observed in the MATRISK study, a prospective multicenter observational study of consecutive iTTP and HUS conducted over 3 years in metropolitan France.

## Methods

### Patients

Participants of this study were included in MATRISK, a prospective multicenter observational study in French Hospitals between June 2014 and July 2017 with follow-up investigations over 6 months (Clinicaltrials.gov Identifier: NCT02134171). Individuals were selected based on age ≥18-year-old, anemia with negative direct antiglobulin test, hemolysis with presence of schistocytes on blood smear, thrombocytopenia (platelet count <150G/L), lack of a secondary cause of TMA that might influence the prognosis, and Imaging data obtained during the first week of hospitalization. Patients gave their informed consent to participate. When they were unable to provide such informed consent because of their clinical status, the consent was first obtained by the patient’s trusted person. The study was approved by an independent ethics committee. The methodology complied with STROBE recommendations (**Supplemental File 1**).

### Clinical data

The following information were recorded as per protocol (**Supplemental File 2**) in each patient at entry in the study: age and gender, any previous history of cardiac or neurological disorder or any recent infection or diarrhea preceding the hospitalization, arterial blood pressure, any clinical manifestation suggestive of infection or diarrhea, cardiac dysfunction such as chest angina, heart failure or cardiogenic shock, any neurological disturbances such as transient ischemic attacks, completed stroke, confusion or coma. Standard biological explorations were assessed. The occurrence of any additional cardiac or neurological symptom over the first 3 days of hospitalization was thereafter systematically recorded. The neurological status of each individual was also assessed using the NIH-stroke scale (NIHSS)^7^ to determine the severity of focal neurological deficits and with the Mini Mental Status Examination (MMSE) score^8,9^, when possible, for evaluating the cognitive status with the modified Rankin Scale for estimating the global severity of disability^10^. ADAMTS13 activity (%) and the level of anti-ADAMTS13 IgG (U/mL) were recorded at admission. Cardiac troponin I was obtained repeatedly the first 3 days with a threshold of 0.20 ng/mL considered as significantly elevated.

### Cerebral Imaging data

Cerebral magnetic resonance imaging (MRI) was obtained within the first 7 days of hospitalization. The following sequences were obtained; 3D-T1 weighted images with millimetric resolution; T2- or FLAIR-weighted images with slices of 5 mm thickness or less; T2*-weighted images (T2* images) of 5 mm thickness for searching old micro or macrobleeds (hypointensities) and recent hemorrhagic lesions with considering the other MRI sequences; diffusion-weighted MRI (DWI) for identifying the most recent ischemic lesions as hyperintense lesions with decreased apparent diffusion coefficient. In addition, Magnetic Resonance Angiography was systematically obtained for assessing the status of large intracranial vessels.

All images were assessed blinded to the status of each individual by LN using a structured assessment and scoring form. When lesions were difficult to classify, the decision was made by consensus (LN and HC). First, on FLAIR images, all types of lesions were assessed, particularly, old infarcts as small cavities with signal identical to that cerebrospinal fluid or focal hyperintensities without signal changes on DWI as well as the presence and extension of white-matter hyperintensities suggestive of the presence of a small vessel disease according to the Fazekas Scale (0=absent; 1=punctate foci; 2=beginning confluence; 3=large confluent areas). On DWI, the presence and number of hyperintense lesions corresponding to recent ischemic lesions was also assessed with their different size (more or less than 20 mm in diameter)^11^. On T2*-weighted images, the presence of old hemorrhagic lesions (as hypointensities) as well as recent hemorrhagic lesions was systematically recorded. In addition, the presence of stenosis or any occlusion of large vessels was assessed on MR angiography.

### Statistical analysis

Data were described using percentage, mean and standard deviation or median values. For each variable, normality was assessed with graphic analysis and using the normal quantile plot with the Shapiro-Wilk test. Univariate analysis was first obtained for comparison between different groups. The comparison was performed based on the Exact Fisher’s test for categorical variables, the Student t test for continuous variables of normal distribution and the Wilcoxon test in the absence of normality. Multivariable binomial logistic models were fit to find the final and best models using the Akaike criterion in order to predict: the occurrence of recent small ischemic lesions on DWI or that of all types of recent cerebral lesions on MRI. This was performed after selection of factors among those with p value <0.2 in the univariate analysis. Only variables with less than 5% of missing values were used in the final analysis. The missing data, if any, were replaced using five replicate imputations based on the chained equation method for multiple imputation (package mice with R). Internal validation of the prediction models was based on the c-index, the Somers’ Dxy rank correlation, a bias-corrected index, obtained after nonparametric bootstrapping with 1000 resamples. The level of significance was set at 0.05 (2-tailed). Statistical analysis was performed using JMP14.3 (SAS) and RStudio, 2022.12.0.

## Results

### Clinical Information

Eighteen French centers participated to the study. Among the 118 patients initially included in the study, 21 patients were subsequently excluded because TMA was related to a clear underlying condition that could have influenced the prognosis. Seventeen patients had no MRI within 7 days after inclusion. Seven additional individuals did not complete the 6-month follow-up. Finally, 73 TMA cases (50 women, 23 men, mean age 46.9±16 years (range: 21-87 years)) were included, either with iTTP when ADAMTS13 activity was <10% and anti-ADAMTS13 antibodies were detectable or ADAMTS13 activity recovered following the acute phase (n=57) or with HUS when ADAMTS13 activity was normal (n=16, STEC-HUS [n=10] or atypical HUS [n=6]) (**Supplemental Figure 1**).

At inclusion or during the first 3 days of hospitalization, 6 (8.2%) patients developed a coma, 15 (20%) an episode of confusion, 9 (12%) presented with focal or generalized epileptic seizures, and 23 (31%) had a focal deficit at neurological examination. A total of 40 (55%) individuals did not present with any of these neurological symptom. Mean MMSE score was 24±7 (range: 4-30), 19 patients (26%) presented with significant cognitive impairment (MMSE score <24) during the first day of hospitalization, 7 (9.5%) of them had with severe cognitive alterations (MMSE score <10). Sixteen patients (22%) reported headache. Clinical presentation did not differ between the two groups of patients except age, which was older in HUS patients. Biological differences were consistent with the diagnosis of patients^12^; iTTP patients had more frequently an elevation of Troponin (**Table 1**).

**Table 1.**
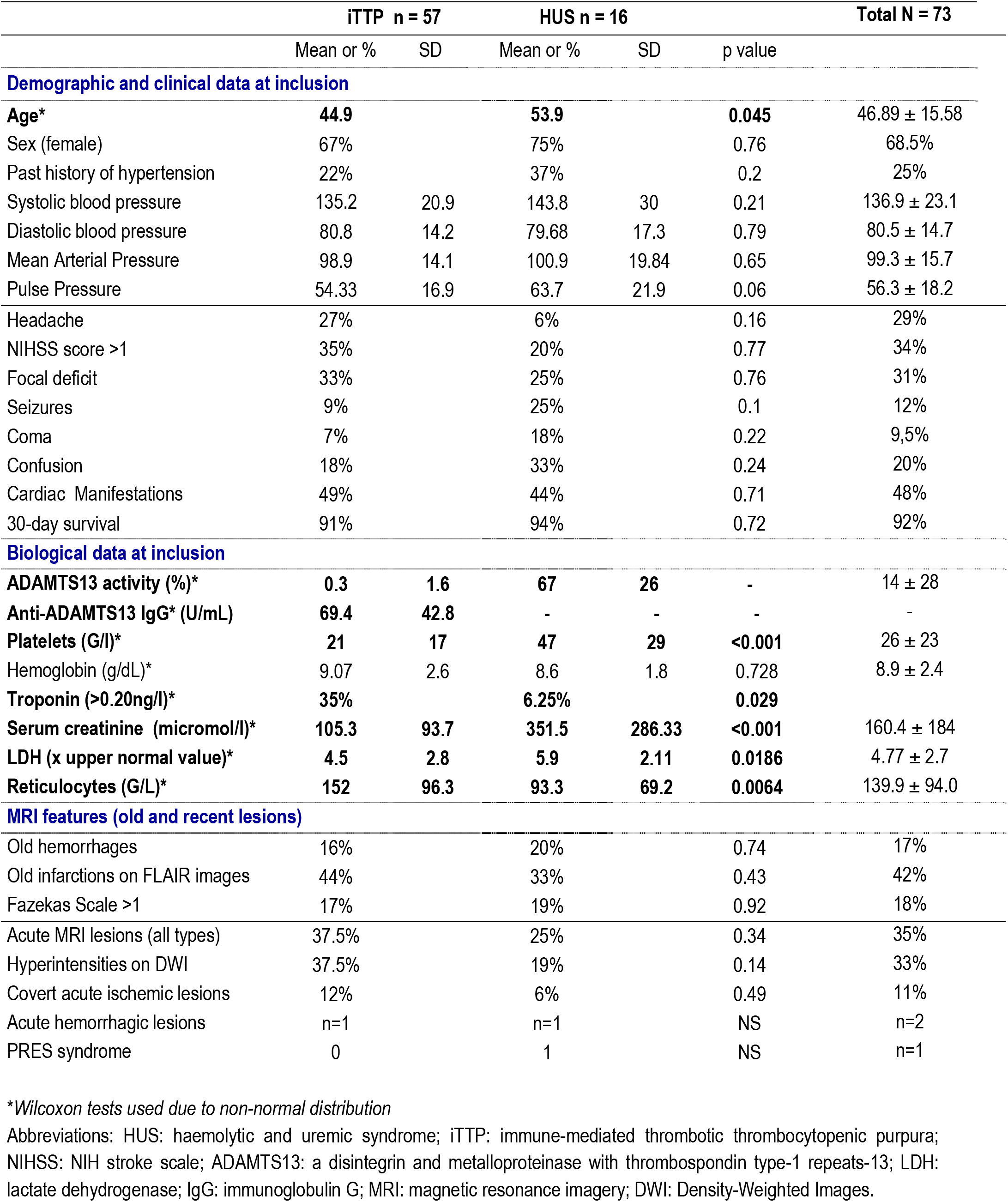
Main clinical, biological and MRI features in the two groups of patients.

|  | iTTP n = 57 |  | HUS n = 16 |  |  | Total N = 73 |
| --- | --- | --- | --- | --- | --- | --- |
|  | Mean or % | SD | Mean or % | SD | p value |  |
| **Demographic and clinical data at inclusion** |  |  |  |  |  |  |
| Age* | 44.9 |  | 53.9 |  | 0.045 | 46.89 ± 15.58 |
| Sex (female) | 67% |  | 75% |  | 0.76 | 68.5% |
| Past history of hypertension | 22% |  | 37% |  | 0.2 | 25% |
| Systolic blood pressure | 135.2 | 20.9 | 143.8 | 30 | 0.21 | 136.9 ± 23.1 |
| Diastolic blood pressure | 80.8 | 14.2 | 79.68 | 17.3 | 0.79 | 80.5 ± 14.7 |
| Mean Arterial Pressure | 98.9 | 14.1 | 100.9 | 19.84 | 0.65 | 99.3 ± 15.7 |
| Pulse Pressure | 54.33 | 16.9 | 63.7 | 21.9 | 0.06 | 56.3 ± 18.2 |
| Headache | 27% |  | 6% |  | 0.16 | 29% |
| NIHSS score >1 | 35% |  | 20% |  | 0.77 | 34% |
| Focal deficit | 33% |  | 25% |  | 0.76 | 31% |
| Seizures | 9% |  | 25% |  | 0.1 | 12% |
| Coma | 7% |  | 18% |  | 0.22 | 9,5% |
| Confusion | 18% |  | 33% |  | 0.24 | 20% |
| Cardiac Manifestations | 49% |  | 44% |  | 0.71 | 48% |
| 30-day survival | 91% |  | 94% |  | 0.72 | 92% |
| **Biological data at inclusion** |  |  |  |  |  |  |
| ADAMTS13 activity (%)* | 0.3 | 1.6 | 67 | 26 | - | 14 ± 28 |
| Anti-ADAMTS13 IgG* (U/mL) | 69.4 | 42.8 | - | - | - | - |
| Platelets (G/l)* | 21 | 17 | 47 | 29 | <0.001 | 26 ± 23 |
| Hemoglobin (g/dL)* | 9.07 | 2.6 | 8.6 | 1.8 | 0.728 | 8.9 ± 2.4 |
| Troponin (>0.20ng/l)* | 35% |  | 6.25% |  | 0.029 |  |
| Serum creatinine (micromol/l)* | 105.3 | 93.7 | 351.5 | 286.33 | <0.001 | 160.4 ± 184 |
| LDH (x upper normal value)* | 4.5 | 2.8 | 5.9 | 2.11 | 0.0186 | 4.77 ± 2.7 |
| Reticulocytes (G/L)* | 152 | 96.3 | 93.3 | 69.2 | 0.0064 | 139.9 ± 94.0 |
| **MRI features (old and recent lesions)** |  |  |  |  |  |  |
| Old hemorrhages | 16% |  | 20% |  | 0.74 | 17% |
| Old infarctions on FLAIR images | 44% |  | 33% |  | 0.43 | 42% |
| Fazekas Scale >1 | 17% |  | 19% |  | 0.92 | 18% |
| Acute MRI lesions (all types) | 37.5% |  | 25% |  | 0.34 | 35% |
| Hyperintensities on DWI | 37.5% |  | 19% |  | 0.14 | 33% |
| Covert acute ischemic lesions | 12% |  | 6% |  | 0.49 | 11% |
| Acute hemorrhagic lesions | n=1 |  | n=1 |  | NS | n=2 |
| PRES syndrome | 0 |  | 1 |  | NS | n=1 |

\*Wilcoxon tests used due to non-normal distribution
Abbreviations: HUS: haemolytic and uremic syndrome; iTTP: immune-mediated thrombotic thrombocytopenic purpura; NIHSS: NIH stroke scale; ADAMTS13: a disintegrin and metalloproteinase with thrombospondin type-1 repeats-13; LDH: lactate dehydrogenase; IgG: immunoglobulin G; MRI: magnetic resonance imagery; DWI: Density-Weighted Images.

Among the 73 patients of the cohort, 62 were treated by therapeutic plasma exchange in association with corticosteroids in 57, and Rituximab in 39. Eculizumab was used in 10 HUS patients. At the end of first 30 days, 67 patients were surviving, as at 3 months. An additional patient died before the 6 month-follow-up. Among survivors, the modified Rankin Scale was recorded in 32 individuals at 6 months; this score was 0 in 26 patients, 1 in 5 patients and 3 in a single subject. MMSE score measured in these subjects varied from 20 to 30 (median = 30 [IQR, 28-30]).

### Imaging results

The main MRI features are summarized in **Table 1**. The presence of old cerebral lesions was detected as focal small infarcts in 42% of patients, micro or macrobleeds in 17% of individuals and white-matter hyperintensities in 18%. Acute ischemic lesions were observed in one third of patients; of note, these lesions were found in the absence of any focal deficit, seizure, confusion or coma (“covert” ischemic lesions) in 11% of cases. On the other hand, seizures, confusion or even coma, were observed in 15 patients without any acute cerebrovascular lesion visualized on their MRI. Among the 24 individuals with acute ischemic lesions detected on MRI, 15 (63%) presented only small hyperintensities on DWI, 4 (16%) only large lesions, and 5 (20%) had both large and small lesions (**Figure 1**). Acute haemorrhagic lesions were detected only in 2 cases, in association with multiple recent ischemic lesions. We observed features highly suggestive of a posterior reversible encephalopathy syndrome (PRES) in only one case with HUS. The analysis did not reveal significant differences in imaging findings between iTTP and HUS patients.

**Figure 1.**
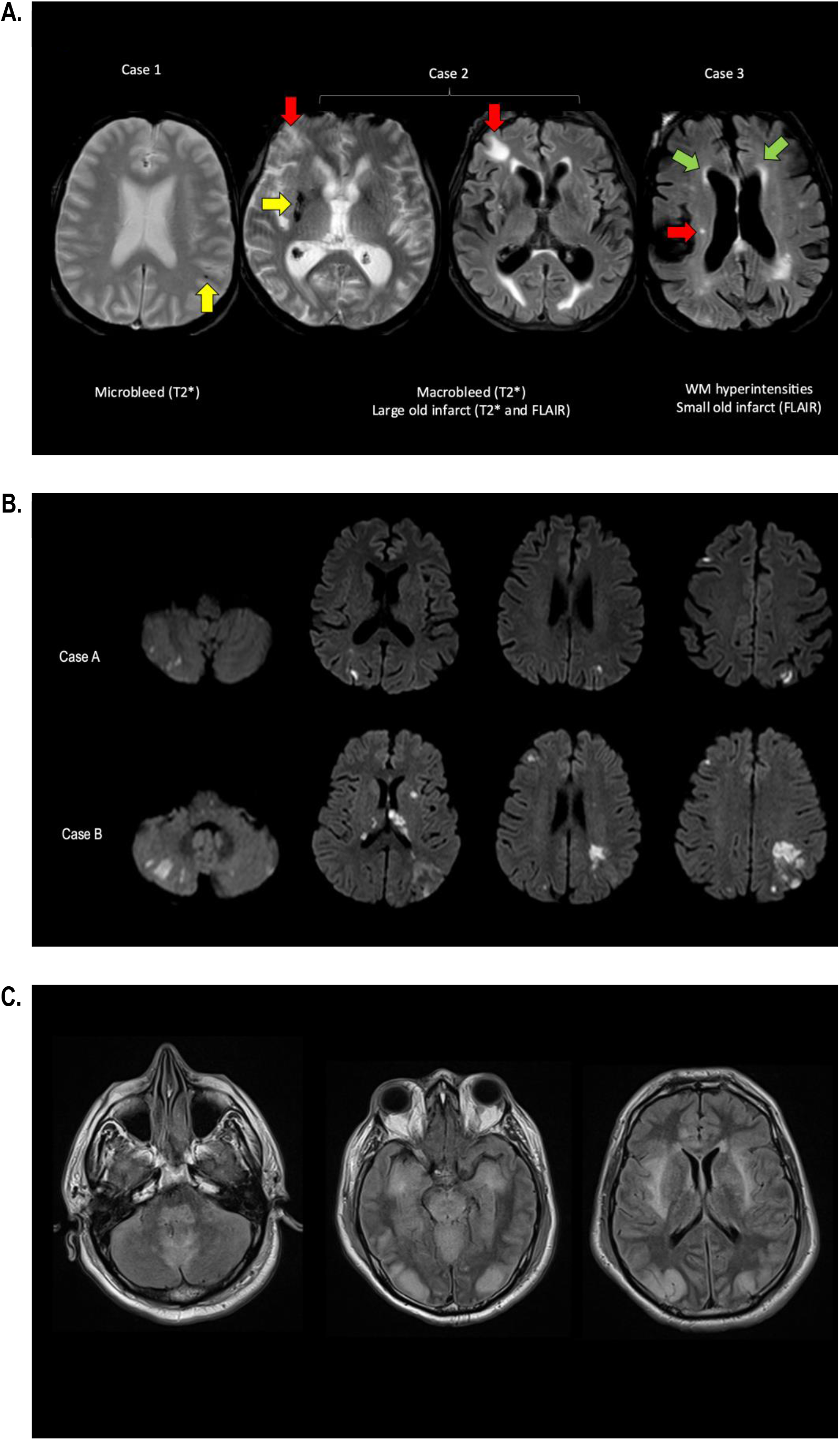
**A)** Old lesions were systematically recorded on T2* images as microbleeds (hypointensity with diameter less than 20 mm) (yellow arrow case 1), macrobleeds (hypointensity of diameter larger than 20 mm, yellow arrow case 2) or old infarcts using FLAIR (Fluid Attenuated Inversion Recovery) images and DWI (Diffusion-Weighted Images) (focal hyperintense lesion with or without cavitation and no signal change on DWI, red arrows case 2 and 3). The extension of white-matter hyperintensities (green arrow, case 3) was also analyzed. **B)** Recent ischemic lesions identified in one third of patients with immune-mediated thrombotic thrombocytopenic purpura and hemolytic and uremic syndrome on diffusion-weighted images (DWI). DWI obtained in two individuals showed the most frequent pattern of ischemic lesions as observed in Case A with multiple recent small ischemic lesions (less than 15 mm in diameter) in superficial cortical areas in as well as in the cerebellum. The pattern observed in case B included both small superficial ischemic lesions and larger recent ischemic lesions either at cortical or cerebellar level. **C)** Atypical lesions observed in the single PRES (posterior reversible encephalopathy syndrome) case selected in the study sample on FLAIR (Fluid Attenuated Inversion Recovery) images and corresponding to cerebral tissue edema related to the disruption of the blood brain barrier.

### Predictors of acute ischemic lesions

We analysed first patients who presented with acute ischemic lesions visualized on DWI in comparison to the others (haemorrhagic cases were included as they also had multiple ischemic lesions). Univariate analysis showed that patients with acute ischemic lesions were older, more frequently hypertensive prior to the hospitalization and had more frequent old cerebral infarcts on their MRI. They also had a higher systolic blood pressure and Pulse Pressure (Systolic-Diastolic Blood Pressure), at time of entry in the study. As expected, they had more frequently neurological deficits and confusion. Finally, 3-month overall survival was also lower in this group. Similar results were obtained when considering the PRES case in the analysis (**Table 2**; **supplementary Table**).

**Table 2.** Univariate analysis of differences between patients with and without acute ischemic lesions on DWI.

|  | Absent n = 47 |  | Present n = 25* |  |  |
| --- | --- | --- | --- | --- | --- |
|  | Mean or % | SD | Mean or % | SD | *p value* |
| Demographic and clinical data at inclusion |  |  |  |  |  |
| Age | 42.90 | 14.02 | 54.17 | 17.28 | 0.0085* |
| Sex (female) | 67% |  | 75% |  | 0.5907 |
| Past history of hypertension | 17% |  | 40% |  | 0.0441 |
| Systolic blood pressure (mm Hg) | 132.09 | 20.34 | 146.24 | 25.98 | 0.0121 |
| Diastolic blood pressure (mm Hg) | 81.38 | 12.81 | 79 | 18.18 | 0.5202 |
| Mean Arterial Pressure (mm Hg) | 98.30 | 13.42 | 101.41 | 19.66 | 0.422 |
| Pulse Pressure (mm Hg) | 50.70 | 16.84 | 67.88 | 16.30 | 0.0001* |
| Cardiac Manifestations | 48% |  | 48% |  | 1 |
| Headache | 21% |  | 25% |  | 0.7667 |
| NIHSS score >0 | 19% |  | 63% |  | 0.0004 |
| Focal déficit | 25% |  | 44% |  | 0.1166 |
| Seizures | 8% |  | 20% |  | 0.4275 |
| Coma | 8.3% |  | 8% |  | 0.960 |
| Confusion | 10% |  | 37.5 |  | 0.0105* |
| 30-day survival | 98% |  | 80% |  | 0.0139 |
| Biological data at inclusion |  |  |  |  |  |
| HUS vs iTTP | 27% |  | 12% |  | 0.2325 |
| ADAMTS13 activity (%) | 16.35 | 32.20 | 5.91 | 18.16 | 0.0904 |
| Anti-ADAMTS13 IgG (U/mL) | 56.81 | 43.48 | 75.20 | 47.36 | 0.1569 |
| Platelets (G/l) | 27.02 | 21.87 | 26.72 | 26.01 | 0.9585 |
| Hemoglobin (g/dL) | 9.21 | 2.60 | 8.49 | 2.21 | 0.2276 |
| Troponin (>0.20ng/l) | 16% |  | 48% |  | 0.0063 |
| Serum creatinine (micromol/l) | 153.06 | 181.08 | 177.70 | 202.87 | 0.6231 |
| LDH (x upper normal value) | 4.64 | 2.69 | 5.15 | 3.04 | 0.514 |
| Reticulocytes (G/L) | 135.67 | 97.97 | 154.92 | 83.06 | 0.4259 |
| MRI features (old lesions) |  |  |  |  |  |
| Old hemorrhages | 15% |  | 22% |  | 0.507 |
| Old infarctions on FLAIR images | 15% |  | 96% |  | <.0001* |
| Fazekas Scale >1 | 10% |  | 33% |  | 0.0246* |
| Stenosis of large arteries on MRA (>1) | 55% |  | 35% |  | 0.177 |

\* One patient with a PRES was excluded from this analysis.
Abbreviations: NIHSS: NIH stroke scale; HUS: haemolytic and uremic syndrome; iTTP: immune-mediated thrombotic thrombocytopenic purpura; ADAMTS13: a disintegrin and metalloproteinase with thrombospondin type-1 repeats-13; LDH: lactate dehydrogenase; IgG: immunoglobulin G; MRI: magnetic resonance imagery; FLAIR: Fluid Attenuated Inversion Recovery; DWI: Density-Weighted Images; Magnetic Resonance Angiography.

A multivariable analysis was performed to identify predictors of acute ischemic lesions (**Table 3**). Two models were selected: the first model was obtained with only demographic, clinical and biological data selected by univariate analysis; the second was obtained with including the MRI features of old lesions. In the first model, three factors were independently associated with the occurrence of recent ischemic lesions on diffusion MRI: the level of pulse pressure, a history of hypertension, and a diagnosis of iTTP (vs HUS). When imaging features were also considered, the best model showed that the 3 significant independent factors predicting the occurrence of acute ischemic lesions at MRI examination were first the presence of old cerebral infarcts on the MRI; second, a high level of pulse pressure; and third, a diagnosis of iTTP (vs HUS). These results did not change when the occurrence of a focal deficit (presumably related to a stroke event) or when the level of systolic blood pressure were forced in the final model (not shown). When considering all types of lesions (i.e., including the case of PRES among cases with acute lesions), the presence of old cerebral infarcts on MRI and the level of blood pulse pressure were still two independent factors predicting the occurrence of acute cerebral lesions. During internal validation, the models retained their excellent discrimination across the bootstrap samples.

**Table 3.** Multivariable logistic regression models with validation tests results *((C-index (AUC) is the concordance index, Dxy is the Somer’s index, R2 is the Nagerkerke R*^*2*^ *index)*

| Term | Estimate | Std Error | ChiSquare | P>ChiSq | Lower 95% | Upper 95% |
| --- | --- | --- | --- | --- | --- | --- |
| Intercept | -6.69 | 1.799 | 13.87 | 0.0002 | -10.22 | -3.17 |
| iTTP vs HUS | 1.55 | 0.581 | 7.1 | 0.0077 | 0.40 | 2.69 |
| Pulse Pressure | 0.09 | 0.025 | 13.24 | 0.0003 | 0.042 | 0.14 |
| Past history of hypertension | -0.98 | 0.40 | 5.97 | 0.0145 | -1.77 | -0.19 |
|  | OR | Lower 95% | Upper 95% | Validation tests (Bootstrap, n = 1000) |  |  |
| Pulse Pressure (per mmHg) | 1.097 | 1.043 | 1.153 | *C-index* |  | 0.848 |
| iTTP vs HUS | 22.20 | 2.26 | 217.16 | *Dxy origin (optimism)* |  | 0.694 (0.034) |
| Past history of hypertension | 7.18 | 1.47 | 34.98 | *R2 origin (optimism)* |  | 0.473 (0.055) |

| Term | Estimate | Std Error | ChiSquare | P>ChiSq | Lower 95% | Upper 95% |
| --- | --- | --- | --- | --- | --- | --- |
| Intercept | -6.20 | 2.24 | 7.67 | 0.0056 | -10.60 | -1.81 |
| Old infarcts on FLAIR images (0) | -2.36 | 0.58 | 16.61 | <.0001 | -3.50 | -1.22 |
| Pulse Pressure | 0.069 | 0.03 | 5.07 | 0.0243 | 0.008 | 0.129 |
| iTTP vs HUS | 1.32 | 0.66 | 3.94 | 0.0473 | 0.016 | 2.63 |
|  | OR | Lower 95% | Upper 95% | Validation tests (Bootstrap, n = 1000) |  |  |
| Old infarcts on FLAIR images | 113.39 | 11.65 | 1103.37 | *C-index* |  | 0.946 |
| Pulse Pressure (per mmHg) | 1.07 | 1.009 | 1.139 | *Dxy origin (optimism)* |  | 0.893 (0.018) |
| iTTP vs HUS | 14.17 | 1.03 | 194.55 | *R2 origin (optimism)* |  | 0.767 (0.045) |

| Term | Estimate | Std Error | ChiSquare | P>ChiSq | Lower 95% | Upper 95% |
| --- | --- | --- | --- | --- | --- | --- |
| Intercept | -3.689 | 1.490 | 6.130 | 0.013 | -7.039 | -3.689 |
| Old infarcts on FLAIR images (0) | -1.990 | 0.436 | 20.790 | <.0001 | -3.004 | -1.990 |
| Pulse Pressure | 0.048 | 0.024 | 4.120 | 0.043 | 0.004 | 0.048 |
|  | OR | Lower 95% | Upper 95% | Validation tests (Bootstrap, n = 1000) |  |  |
| Old infarcts on FLAIR images | 53.471 | 11.654 | 406.351 | *C-index* |  | 0.930 |
| Pulse Pressure | 1.049 | 1.004 | 1.105 | *Dxy origin (optimism)* |  | 0.860 (0.030) |
|  |  |  |  | *R2 origin (optimism)* |  | 0.701 (0.055) |

Abbreviations: HUS: haemolytic and uremic syndrome; iTTP: immune-mediated thrombotic thrombocytopenic purpura; FLAIR: Fluid Attenuated Inversion Recovery; OR: Odd-Ratio.

## Discussion

One third of individuals with either iTTP or HUS presented here with acute cerebral ischemic lesions on MRI at time of their hospitalization. These patients were more than 10 years older, and presented more frequently with confusion, seizure and focal deficits than individuals in the rest of the sample. Moreover, their survival was more often compromised despite therapeutic measures. Altogether, these data suggest that the occurrence of these cerebral lesions on MRI at the acute phase of iTTP or HUS has a poor prognostic value. Of note, simultaneous acute hemorrhagic lesions were rare events only observed in two individuals in presence of ischemic lesions. This would rather support the use of caplacizumab, the first anti-von Willebrand factor nanobody drug, as a potential frontline treatment in cases of suspected iTTP^13^. On the other hand, seizures, confusion or even coma, could be observed without any acute cerebrovascular lesion visualized on MRI. This further supports that mechanisms distinct from vascular insults can also lead to severe neurological manifestations at the acute phase of iTTP and HUS. These features might result from brain electrical dysfunction, tissue inflammation, ionic disturbances or perivascular gliosis already reported in these severe conditions^14^. In 15% of cases, we observed focal neurological deficits in association with old infarcts or hemorrhages, suggesting that a significant proportion of patients with iTTP or HUS presented with old cerebrovascular lesions prior their acute decompensation. Whether these lesions resulted from previous exposure to vascular risk factors such as hypertension, from a persistent defect in ADAMTS13^15^, from aging effects or were secondary to some previous cerebrovascular manifestations of TMA remains undetermined.

Here, neurological manifestations were globally less frequent than previously described in iTTP, whose frequency varied between 50 and 92%. The single PRES case in this study for both iTTP and HUS also contrasts with the high frequency of PRES repeatedly observed previous studies^16-17^. The frequency of confusion, seizures, stupor or coma was also lower than that observed in the largest series of TMA^18^. Recruitment bias, differences in the management and treatment, the wider inclusion criteria as well as the lack of measures of ADAMTS13 activity for categorizing patients in some studies, might explain this discrepancy. Interestingly, one patient out of 10 in our study developed incident ischemic lesions without any clinical manifestation or symptom, suggesting that “covert” infarcts could also occur during iTTP or HUS and might have been often unrecognized in previous studies. Cognitive impairment might develop as a possible consequence of accumulated vascular lesions in the brain. In this study, one fourth of patients presented with cognitive alterations during the first days of hospitalization. Among the 32 survivors evaluated at 6-month follow-up, only one presented with significant cognitive and/or motor impairment. Therefore, although we cannot raise definitive conclusions, it is very likely that accumulation of cerebral infarcts or hemorrhages could have a significant impact on cognitive performances in such circumstances.

An important result of this study is that a history of hypertension, an increased level of pulse pressure, i.e., a large difference between systolic and diastolic blood pressure, as well as the diagnosis of iTTP (vs HUS), were found to represent strong and independent predictors of acute ischemic cerebral lesions. Moreover, when imaging data were considered in the multivariable model analysis, the presence of old cerebral infarcts on FLAIR images in association with the level of pulse pressure were explaining alone the occurrence of all types of acute cerebrovascular lesions, and with the additional presence of iTTP diagnosis, the occurrence of acute ischemic lesions. We believe these results might have pathophysiological correlates. There is accumulating data suggesting that the vascular endothelium would be the primary site of injury during TMA. In presence of a severe ADAMTS13 deficiency as observed in iTTP, accumulation of von Willebrand factor multimers on endothelium and in the blood circulation is presumably promoting platelet aggregation. However, additional factors including increased shear stress, toxins or dysregulation of the complement cascade as observed in the different categories of TMA have been identified as a potential second hit to precipitate the occurrence of vascular thrombosis^19^. Experimentally, increasing pulse pressure was reported to induce mechanical stress on cerebral vascular endothelial cells and subsequently endothelial dysfunction from moderate alterations of tight junctions to severe blood brain barrier alterations. From our results, we could hypothesize that the combination of severe ADAMTS13 deficiency in iTTP, high pulse pressure directly acting on the endothelium of the cerebral microvasculature, as well as preexisting structural changes at the same level, dramatically increase the risk of ischemic manifestations in this setting. Additional studies are essential to confirm this hypothesis.

We are aware of important limitations in this study. Although iTTP and HUS are rare conditions, the size of our sample was relatively limited which might have reduced the statistical power and prevented the identification of other key factors influencing the risk of acute cerebrovascular lesions. We also made the choice of not including TMA related to very specific conditions. Thus, our data could not be representative of all categories of TMA. The study was also multicentric, and MRI data were not obtained at the exact same time and using the same scanner in all patients. This might be a source of variable sensitivity in the detection of all acute lesions among patients. Moreover, enrollment of iTTP patients occurred before the recent development of caplacizumab, which further improves their therapeutic management, raising concerns about the external validity of our results. Conversely, this study had also important strengths. MRI results were obtained in individuals who were highly selected based on a large work-up and using an extensive biological assessment including ADAMTS13 measurement. Imaging data were evaluated centrally by two expert neurologists and by consensus in difficult situations. MRI data at entry in the study were complete and allowed assessing not only old infarcts and hemorrhages but also the most recent lesions. Finally, the statistical model had a strong internal validation and allowed obtaining a solid prediction using a limited number of factors with AUC higher than 0.8 in all models.

We believe our results are important to improve the management of patients with iTTP and HUS for two reasons. First, they illustrate the importance of systematic brain MRI examination from the moment of hospitalization not only to detect all ischemic lesions occurring during patients’ management but also to better identify patients at the highest risk of neurological worsening. Second, they suggest that the level of pulse pressure, even in the absence of malignant arterial hypertension, should be considered as an important and independent predictor of ischemic manifestation of TMA and possibly, as a potential target for improving the acute management of these patients.

## Data Availability

Data are available on reasonable request.

## Acknowledgments

The sponsor was Assistance Publique Hôpitaux de Paris (APHP). Patients were recruited with the help of the members of the Reference Center for Thrombotic Microangiopathies (CNR-MAT) (listed in the appendix). We thank S. Thouzeau and S. Capdenat (Laboratoire d’Hématologie, Hôpital Lariboisière, AP-HP, Paris) for technical assistance and URCEST for logistical help. This work was funded by a grant from the French Ministry of Health (Projet Hospitalier de Recherche Clinique 2012; AOM12259). This work was also supported by the National Plan for Rare Diseases of the French Ministry of Health (Direction Générale de l’Offre de Soin (DGOS)). Data on which this article is based can be made available upon reasonable request to the sponsor.

## Disclosures

E.M. declares having received lecture fees from Sanofi and Takeda; A.V. is member of the Clinical Advisory Board for Sanofi and Takeda; P.C. is member of the Clinical Advisory Board for Alexion, Sanofi and Takeda; H.C. is member of an Advisory Board of Biogen and Hovid. Y.B., F.P., Pa.P. and A.W. are members of the clinical advisory board for Sanofi. The other authors declared no conflict of interest.

## Figure legend

**Supplemental Figure 1**. Study flow chart.

*Malignant hypertension [n = 5], cancer [n = 3], HELLP syndrome [n = 3], delivery hemorrhage or fetal death [n = 3], myositis [n = 1], pulmonary hypertension [n = 1], drug or toxic [n = 3], infection with *Capnocytophaga canimorsus* [n = 1], and antiphospholipid syndrome [n = 1].

### Appendix

#### The members of the Reference Center for Thrombotic Microangiopathies (CNR-MAT) are

Augusto Jean-François (Service de Néphrologie, dialyse et transplantation; CHU Larrey, Angers); Azoulay Elie (Service de Réanimation Médicale, Hôpital Saint-Louis, Paris); Barbay Virginie (Laboratoire d’Hématologie, CHU Charles Nicolle, Rouen); Benhamou Ygal (Service de Médecine Interne, CHU Charles Nicolle, Rouen); Charasse Christophe (Service de Néphrologie, Centre Hospitalier de Saint-Brieuc); Charvet-Rumpler Anne (Service d’Hématologie, CHU de Dijon); Chauveau Dominique, Ribes Davis (Service de Néphrologie et Immunologie Clinique, CHU Rangueil, Toulouse); Choukroun Gabriel (Service de Néphrologie, Hôpital Sud, Amiens); Coindre Jean-Philippe (Service de Néphrologie, CH Le Mans); Coppo Paul (Service d’Hématologie, Hôpital Saint-Antoine, Paris); Delmas Yahsou (Service de Néphrologie, CHU de Bordeaux, Bordeaux); Kwon Theresa (Service de Néphrologie Pédiatrique, Hôpital Robert Debré, Paris); Salanoubat Célia (Service d’Hématologie, Hôpital Sud-Francilien, Corbeil-Essonnes); Dossier Antoine (Service de Néphrologie, Hôpital Bichat, Paris); Fain Olivier (Service de Médecine Interne, Hôpital Saint-Antoine, Paris); Ville Simon (Service de Néphrologie, CHU Hôtel-Dieu, Nantes); Frémeaux-Bacchi Véronique (Laboratoire d’Immunologie, Hôpital Européen Georges Pompidou, Paris); Galicier Lionel (Service d’Immunopathologie, Hôpital Saint-Louis, Paris); Grangé Steven (Service de Réanimation Médicale, CHU Charles Nicolle, Rouen); Guidet Bertrand (Service de Réanimation Médicale, Hôpital Saint-Antoine, Paris); Halimi Jean-Michel (Service de Néphrologie Pédiatrique, Hôpital Bretonneau, Tours); Hamidou Mohamed, Neel Antoine (Service de Médecine Interne, Hôtel-Dieu, Nantes); Fornecker Luc-Matthieu (service d’Oncologie et d’Hématologie, Hôpital de Hautepierre, Strasbourg); Hié Miguel (Service de Médecine Interne, Groupe Hospitalier Pitié-Salpétrière, Paris); Jacobs Frédéric (Service de Réanimation Médicale, Hôpital Antoine Béclère, Clamart); Joly Bérangère (Service d’Hématologie Biologique, Hôpital Lariboisière, Paris); Kanouni Tarik (Unité d’Hémaphrèse, Service d’Hématologie, CHU de Montpellier); Kaplanski Gilles (Service de Médecine Interne, Hôpital la Conception, Marseille); Rieu Claire (Hôpital d’Estaing, Service de Médecine Interne, Clermont-Ferrand); Le Guern Véronique (Unité d’Hémaphérèse, Service de Médecine Interne, Hôpital Cochin, Paris); Moulin Bruno (Service de Néphrologie, Hôpital Civil, Strasbourg); Rebibou Jean-Michel (Service de Néphrologie, CHU de Dijon); Ojeda Uribe Mario (Service d’Hématologie, Hôpital Emile Muller, Mulhouse); Parquet Nathalie (Unité de Clinique Transfusionnelle, Hôpital Cochin, Paris); Pène Frédéric (Service de Réanimation Médicale, Hôpital Cochin, Paris); Perez Pierre (Service de Réanimation polyvalente, CHU de Nancy); Poullin Pascale (Service d’hémaphérèse et d’autotransfusion, Hôpital la Conception, Marseille); Marie Manon (Service de Néphrologie, CHU Lyon-Sud, Lyon); Presne Claire (Service de Néphrologie, Hôpital Nord, Amiens); Provôt François (Service de Néphrologie, Hôpital Albert Calmette, Lille); Mesnard Laurent (Urgences Néphrologiques et Transplantation Rénale, Hôpital Tenon, Paris); Saheb Samir (Unité d’Hémaphérèse, Hôpital la Pitié-Salpétrière, Paris); Seguin Amélie (Service de Réanimation Médicale, CHU Hôtel-Dieu, Nantes); Servais Aude (Service de Néphrologie, CHU Necker-Enfants Malades); Stépanian Alain (Laboratoire d’Hématologie, Hôpital Lariboisière, Paris); Veyradier Agnès (Service d’Hématologie Biologique, Hôpital Lariboisière, Paris); Vigneau Cécile (Service de Néphrologie, Hôpital Pontchaillou, Rennes); Wynckel Alain (Service de Néphrologie, Hôpital Maison Blanche, Reims); Zunic Patricia (Service d’Hématologie, Groupe Hospitalier Sud-Réunion, la Réunion).

## Notes

### Clinical Trial

NCT02134171

### Author Declarations

Délégation-la Recherche Clinique d’Ile-de-France(DRCI), Hôpital Saint-Louis, Paris, France

## References

1. George JN, Nester CM. Syndromes of thrombotic microangiopathy. The New England journal of medicine 2014; 371(7): 654–66.

2. DeYoung V, Singh K, Kretz CA. Mechanisms of ADAMTS13 regulation. Journal of thrombosis and haemostasis : JTH 2022; 20(12): 2722–2732.

3. Coppo P, Bubenheim M, Azoulay E, Galicier L, Malot S, Bige N et al. A regimen with caplacizumab, immunosuppression, and plasma exchange prevents unfavorable outcomes in immune-mediated TTP. Blood 2021; 137(6): 733–742.

4. Travert B, Rafat C, Mariani P, Cointe A, Dossier A, Coppo P et al. Shiga Toxin-Associated Hemolytic Uremic Syndrome: Specificities of Adult Patients and Implications for Critical Care Management. Toxins (Basel) 2021; 13(5).

5. Fakhouri F, Loirat C. Anticomplement Treatment in Atypical and Typical Hemolytic Uremic Syndrome. Semin Hematol 2018; 55(3): 150–158.

6. Jia Yu JB, Eli Salzberg, Aria Wei, Gloria Gerber, Xiang-Zuo Pan, Alison R. Moliterno, MIchael B. Streiff, Peggy Kraus, Claire Logue, Jennifer Yui, Rakhi P.P. Naik, Hira Latif, Sophie M. Lanzkron, Evan M. Braunstein, Robert Brodsky, Michael R. DeBaun, Doris Lin, Shruti Chaturvedi. Blood 2022.

7. Adams H, Davis P, Leira E, Chang K, Bendixen B, Clarke W et al. Baseline NIH Stroke Scale score strongly predicts outcome after stroke: a report of the Trial of Org 10172 in Acute Stroke Treatment (TOAST). In: Neurology, 1999. pp 126–131.

8. Cockrell JR, Folstein MF. Mini-Mental State Examination (MMSE). Psychopharmacology bulletin 1988; 24(4): 689–92.

9. Biundo R, Weis L, Bostantjopoulou S, Stefanova E, Falup-Pecurariu C, Kramberger MG et al. MMSE and MoCA in Parkinson’s disease and dementia with Lewy bodies: a multicenter 1-year follow-up study. Journal of neural transmission 2016; 123(4): 431–8.

10. Quinn TJ, Dawson J, Walters MR, Lees KR. Reliability of the modified Rankin Scale. Stroke; a journal of cerebral circulation 2007; 38(11): e144. author reply e145.

11. Wardlaw JM, Smith EE, Biessels GJ, Cordonnier C, Fazekas F, Frayne R et al. Neuroimaging standards for research into small vessel disease and its contribution to ageing and neurodegeneration. The Lancet Neurology 2013; 12(8): 822–838.

12. Coppo P, Schwarzinger M, Buffet M, Wynckel A, Clabault K, Presne C et al. Predictive features of severe acquired ADAMTS13 deficiency in idiopathic thrombotic microangiopathies: the French TMA reference center experience. PloS one 2010; 5(4): e10208.

13. Schofield J, Shaw RJ, Lester W, Thomas W, Toh CH, Dutt T. Intracranial hemorrhage in immune thrombotic thrombocytopenic purpura treated with caplacizumab. Journal of thrombosis and haemostasis : JTH 2021; 19(8): 1922–1925.

14. Weil EL, Rabinstein AA. Neurological manifestations of thrombotic microangiopathy syndromes in adult patients. Journal of thrombosis and thrombolysis 2021; 51(4): 1163–1169.

15. Upreti H, Kasmani J, Dane K, Braunstein EM, Streiff MB, Shanbhag S, Moliterno AR, Sperati CJ, Gottesman RF, Brodsky RA, Kickler TS, Chaturvedi S. Reduced ADAMTS13 activity during TTP remission is associated with stroke in TTP survivors. Blood 2019;134(13):1037–1045.

16. Meloni G, Proia A, Antonini G, De Lena C, Guerrisi V, Capria S et al. Thrombotic thrombocytopenic purpura: prospective neurologic, neuroimaging and neurophysiologic evaluation. Haematologica 2001; 86(11): 1194–9.

17. Burrus TM, Wijdicks EF, Rabinstein AA. Brain lesions are most often reversible in acute thrombotic thrombocytopenic purpura. Neurology 2009; 73(1): 66–70.

18. de Castro JTS, Appenzeller S, Colella MP, Yamaguti-Hayakawa G, Paula EV, Annichinno-Bizzachi J et al. Neurological manifestations in thrombotic microangiopathy: Imaging features, risk factors and clinical course. PloS one 2022; 17(9): e0272290.

19. Mathew RO, Nayer A, Asif A. The endothelium as the common denominator in malignant hypertension and thrombotic microangiopathy. J Am Soc Hypertens 2016; 10(4): 352–9.

